# The role of social drivers of health in communication abilities of autistic adolescents and young adults

**DOI:** 10.1101/2024.06.17.24309053

**Authors:** Teresa Girolamo, Alicia Escobedo, Lindsay Butler, Caroline A. Larson, Iván Campos, Kyle Greene-Pendelton

**Author notes:** Consolidated School District of New Britain (Kyle Greene-Pendelton). **Corresponding author**: Teresa Girolamo, School of Speech, Language, and Hearing Sciences, 5201 Campanile Mall, San Diego, CA, 92103.

## Abstract

Despite their relevance to outcomes in autism, little is known about how social drivers of health affect communication, especially in transition-aged autistic adolescents and young adults with structural language impairment. This knowledge gap limits our understanding of developmental trajectories and the ability to develop supports. This cross-sectional study examined the role of social drivers of health in communication abilities of minoritized autistic individuals ages 13 to 30. Participants (*N* = 73) completed language, nonverbal cognitive assessments, and social drivers of health (sense of community, unmet services, barriers to services) measures. Data were analyzed descriptively and using mixed-effects modeling. More unmet service needs, more barriers to services, and lower sense of community were associated with greater social communication impairment. In turn, both unmet service needs and barriers to services were negatively associated with functional communication. In regression modeling, language scores contributed to functional communication, and sense of community to social communication impairment. Findings support the relevance of language and social drivers of health in communication. Future work should focus on possible bidirectional relationships between these variables and explore and real-world translation.

**Lay Abstract:** Where people live, work, and spend their time is important. Environments can have more or less services or differ in how much they help people feel like they belong to their community. These parts of the environment are called social drivers of health. Social drivers of health are important for outcomes in autism, but we do not know much about them in racially and ethnically minoritized autistic teens or young adults. We recruited 73 minoritized autistic teens and young adults (ages 13 to 30 years) and 52 caregivers to our study. Autistic teens and young adults did language and NVIQ tests on Zoom. Autistic teens, young adults, and caregivers also answered questionnaires. Sense of community was important for social communication impairment, and language was important for real-world communication. These findings tell us two things. First, thinking about how to create supportive communication environments for autistic teens and adults is important. Second, understanding how social drivers of health shape outcomes is important. In the future, we should focus on how improving environments can help minoritized autistic teens and adults meet their communication goals.

---

Autistic individuals often face poor adaptive, employment, mental and physical health, and social outcomes in adulthood (Mason et al., 2021). In the U.S., the transition to adulthood is a vulnerable period, coinciding with the loss of child- and school-based services (“Individuals with Disabilities Education Improvement Act [IDEIA],” 2004; Roux et al., 2015). Despite growing attention to adult outcomes (Howlin & Magiati, 2017), understanding remains limited, especially considering the systematic exclusion of racially and ethnically minoritized communities in autism research (Maye et al., 2021). Recent studies reveal racial disparities in health outcomes. For example, Medicaid data—a joint federal and state program providing medical and health services to people with limited income (U.S. Congress, 1965)—show higher rates of cardiovascular disease among Black and Hispanic autistic adults when data are disaggregated (Schott et al., 2022). Other research highlights disparities in hospitalization and diagnosis of mental health conditions (Ames et al., 2022; Rast et al., 2023) This heterogeneity has implications for examining other outcomes in the transition to adulthood.

Person-environment fit is increasingly recognized as central to autism outcomes (Lai et al., 2020), aligning with a social-ecological model where individual-environment interactions shape development (Bronfenbrenner, 1977). Social drivers of health, or non-medical factors like social networks, economic stability, and physical setting, play a key role (Centers for Disease Control and Prevention, 2021; Commission on Social Determinants of Health, 2008). Yet there is limited attention to social drivers of health in autism research, particularly beyond childhood (Schendel et al., 2022). Most research on transition-aged autistic individuals focuses on family-level factors (e.g., income) and overlooks interpersonal- or community-level factors like social inclusion (Anderson et al., 2018a).

Meanwhile, there is a robust literature on the importance of language skills in autism outcomes (Brignell et al., 2018; Magiati et al., 2014). Language is central to communication, which is a human right (McLeod, 2018; United Nations, 1948), and is often impaired. Over 60% of autistic individuals have structural language impairment (LI; Boucher, 2012), or difficulty with grammatical aspects of language like morphology and syntax (Schaeffer et al., 2023). Structural LI is tied to adult communication abilities, which influence educational, occupational, and social outcomes (Johnson et al., 2010). Understanding communication in autism thus requires attention to both linguistic heterogeneity and social drivers of health.

## Contextualizing Communication in Autism

Communication abilities in autistic adolescents and adults vary widely (Anderson et al., 2014; Billstedt et al., 2007; Gillespie-Lynch et al., 2012; Howlin et al., 2013; Levy & Perry, 2011), in part due to differences in measurement. Functional communication refers to individual ability to meet everyday communicative demands using verbal or written language (Sparrow et al., 2016). Social communication impairment reflects difficulties in social communication and interaction (Constantino, 2012) consistent with a social-ecological model (Bronfenbrenner, 1977). Each frames communication differently. For example, prior work shows functional communication scores in autistic adolescents and adults often fell ≥ 2 *SD* below the mean, regardless of cognitive ability (Farley et al., 2009; Matthews et al., 2015). Some autistic adults (ages 22-46) with low functional communication scores reported meaningful social inclusion (Farley et al., 2009), while others face bias in interactions from nonautistic peers – even when their communication is effective (Crompton et al., 2020a; 2020b; Morrison et al., 2020a). These findings underscore the role of context.

Two key limitations hinder understanding communication in transition-aged autistic individuals. First, studies often overlook linguistic heterogeneity. Autism can co-occur with LI and intellectual disability (American Psychiatric Association, 2013), but language and intelligence (IQ) can dissociate (Manenti et al., 2024; Silleresi et al., 2020; World Health Organization [WHO], 2022). Detecting this variation depends upon multi-domain assessment (Kover & Abbeduto, 2023). For instance, Bal et al. (2019) found links between language level and social communication impairment in autistic young adults, but language was based on a single caregiver-report item. In contrast, Johnson et al. (2010) directly assessed multiple language domains in autistic and nonautistic young adults with a history of LI (ages 24-26 years) and found varied correlations between language measures and functional communication scores (*r* = .31-.88). Relatedly, verbal IQ (VIQ) often underestimates ability in autistic youth, especially those with LI (Grondhuis et al., 2018). Findings linking IQ to communication in autistic adolescents and adults are mixed (Lord et al., 2020; Matthews et al., 2015; Tamm et al., 2022), motivating attention in assessment.

Second, social drivers of health are underexamined in relation to disparities (Braveman & Gottlieb, 2014). Practices, such as default comparison of white to minoritized participants (e.g., McCauley et al., 2020), risk perpetuating monolithic stereotypes (Plaut, 2010). U.S. federal policies have shaped how race and disability intersect – including in service systems for autistic adolescents and young adults (Powell, 2012; Turnbull III et al., 2006) – and reflected external factors rather than any traits of minoritized autistic individuals themselves (Annamma et al., 2013). Understanding communication for minoritized autistic adolescents and young adults thus requires attention to interpersonal- and community-level factors (Anderson et al., 2018a).

Unmet service needs and barriers are key social drivers for transition-aged autistic individuals within the U.S. (Burke et al., 2024; Eilenberg et al., 2019; Shattuck et al., 2020). Young adults often report more unmet needs than youth (Anderson et al., 2018b; Laxman et al., 2019; Turcotte et al., 2016), especially for communication-related supports (Ishler et al., 2023; Schott et al., 2021). These needs may increase prior to high school exit (Taylor & Henninger, 2015). Barriers, such as cost, location, quality of services, and provider communication challenges, also may increase with age and persist into young adulthood (Anderson & Butt, 2018; Cummins et al., 2020; Doherty et al., 2022).

A third social driver of health is sense of community, or sense of belonging and feeling supported within a community (McMillan & Chavis, 1986). It is tied to community participation and adaptive behavior (Cameron et al., 2022; Mahmoudi Farahani, 2016; Talò et al., 2014; Zidrou et al., 2021) but may be limited by structural constraints – especially among marginalized groups (Littman, 2022). Supportive environments promote community participation for autistic youth and adolescents (Chen et al., 2023; Tobin et al., 2014), whereas a lack of community understanding about autism can reduce communication opportunities (Billstedt et al., 2010; Morrison et al., 2020b). To this end, U.S. autistic young adults (ages 20-25 years) report greater social isolation than nonautistic peers with disabilities (Orsmond et al., 2013).

## The Current Study

This report examines communication abilities in U.S.-based minoritized autistic adolescents and young adults, focusing on social communication impairment and functional communication. Given heterogeneity within minoritized communities (Plaut, 2010), the premise of this paper is that systematic exclusion in research motivates purposeful inclusion of these individuals to help produce broadly applicable findings (Maye et al., 2021). The focus is on to what extent individual differences, social drivers of health, and communication relate to one another. Specifically, this study asked:

1. Does LI status differentiate groups in functional communication and social communication impairment scores and social drivers of health (sense of community, unmet service needs, and barriers to having service needs met), given NVIQ band?
2. To what extent do individual differences and social drivers of health contribute to differences in functional communication and social communication impairment scores? In contrast to most prior work, this study directly assessed language across domains and

NVIQ, which enhanced our confidence in our findings (Grondhuis et al., 2018). In using norm-referenced measures, this approach also provides information that is readily interpretable and aligns with clinical practice patterns for service eligibility in the U.S. (“Americans with Disabilities Act of 1990,” 2008; “Rehabilitation Act of 1973,” 1973; Selin et al., 2022). Last, in using person-centered measures, this study provides new, expert-informed information on social drivers of health to help build the evidence base on services (Burns et al., 2011).

## Method

This study received institutional board approval and followed all ethical guidelines. Our team also used a community-based participatory approach (Wallerstein & Duran, 2006), which was tailored to a localized context in terms of providing accessible partnership and avoiding tokenization, such as by assuming partners wanted to be defined on the perceptions of *others* about race and disability (Girolamo et al., 2024). Partners chose their role and opted to join the team at all study stages, including co-development of the research questions and study design (methods, outcome measures, interpretation of the data), and dissemination (e.g., community-driven workshops). While the research team included diverse individuals with lived, personal, and professional experiences pertaining to autism, including autistic people from the communities included in this study, team members did not know participants in this study.

## Participants

Selection criteria were: (a) racially minoritized, ethnically minoritized, or both racially and ethnically minoritized according to U.S. Census guidelines (Office of Management and Budget, 1997), which included: American Indian or Alaska Native, Asian, Black or African American, Native Hawai’ian or Pacific Islander, multiracial, and other for race and Hispanic/Latine for ethnicity, with the option to select multiple options for race and to write in options; (b) formal clinical diagnosis of autism, per requirements for inclusion in recruitment sources (e.g., community organizations providing services to autistic individuals), and confirmation using the Social Responsiveness Scale-2^nd^ Ed. (SRS-2; Constantino, 2012) and expert clinical judgment; (c) ages 13 to 30, coinciding with when transition planning begins in some states within the U.S. and 10 years post-federal eligibility for special education services (“Every Student Succeeds Act,” 2015; “IDEIA,” 2004); (d) proficiency in English per self-report during screening, as assessments were in English; (e) adequate hearing and vision thresholds for responding to audiovisual stimuli on a computer screen, and; (f) use of primarily spoken language to communicate, as study activities required oral responses. Participants could be of any sex assigned at birth and gender, with the ability to self-report options.

## Procedures

The research team recruited participants in a multi-step process: (a) sharing virtual flyers with information about the study with organizations serving autistic individuals, (b) providing personalized consultation about the study to individuals and families about the study by phone, Zoom, or email, (c) obtaining informed consent using a dynamic process, and (d) collecting data. In this process, an examiner asked participants and caregivers about communication strategies, preferred modalities, and different communicative acts the participant might use to convey emotions or feelings, such as agreement or needing a break. The team used this information during informed consent and assessment. Participants provided informed consent if they were their own legal guardian. Caregivers provided informed consent if they were the legal guardian of participants, and participants provided assent. In all cases, the examiner went over the consent form line by line with participants (and caregivers, as appropriate), discussing concepts like “consent” and “confidentiality.” The examiner also reviewed who participants could call if they had questions or concerns and options for sharing data (e.g., de-identified scores, transcripts, and/or recordings). Throughout this process, the examiner administered verbal checks and encouraged participants to ask questions (e.g., “Can you stop being in the study at any time?” or “If you stop being in the study, are there any bad consequences?”). If participants provided assent, depending on the communication profiles shared with the research team during consultation, the examiner asked tailored questions (e.g., “Can I tell other people you are in my study?”, “Do you want to try the activities?”). Recruitment and data collection took place remotely on HIPAA-compliant Zoom. The first author administered a behavioral assessment protocol to participants and caregivers at their convenience using test developer guidance on remote assessment for measures of language and NVIQ (Pearson, 2023). Participants and caregivers completed questionnaires.

## Measures

### Autism Traits

Autism traits were assessed using SRS-2 caregiver and self-report forms for students and adults (Constantino, 2012). Respondents indicate the frequency of 65 items on a four-point scale, yielding an overall *t*-score. *T*-scores of ≤ 59 indicate sub-clinical, 60 to 65 mild, 66 to 76 moderate, and >76 high levels of autism traits. Formal diagnosis, SRS-2 *t*-scores, and expert clinical judgment were triangulated to determine autism traits. Overall *t*-scores of caregiver student forms, caregiver adult forms, and adult self-report forms did not significantly differ; see Supplementary Table 1.

### Language Skills

Participants completed a battery of normed assessments across linguistic domains: semantics, morphology, syntax, and phonology. Expressive language and receptive language were assessed by the Clinical Evaluation Language Fundamentals-5^th^ Ed. (CELF-5) Expressive Language Index and Receptive Language Index (*M* = 100, *SD* = 15; Wiig et al., 2013). For those over age 21 (*n* =25), CELF-5 age 21 norms were used per prior studies of adults ages 18 to 49 (Botting, 2020; Clegg et al., 2021; Fidler et al., 2011). Receptive and expressive vocabulary were assessed by the Peabody Picture Vocabulary Test-5^th^ Ed. (PPVT-5; Dunn, 2019) and Expressive Vocabulary Test-3^rd^ Ed. (EVT-3; *M* = 100, *SD* = 15; Williams, 2019). Phonological working memory was assessed by percent accuracy on the Syllable Repetition Task (SRT), a measure of nonword repetition (*M* = 92, *SD* = 5.9 in six-year-old autistic children with FSIQ ≥ 70; Shriberg et al., 2009; Shriberg & Mabie, 2017). LI was defined as ≤ −1.25 *SD* on ≥2 measures: CELF-5 Expressive Language Index, CELF-5 Receptive Language Index, PPVT-5 standard, EVT-3 standard score, or SRT overall accuracy. This cutoff aligns with epidemiological criteria for LI in nonautistic U.S.-based youth (Tomblin et al., 1997). While more stringent than the Ottawa study cutoff of −1 *SD* on local norms for a receptive vocabulary measure or omnibus language measure, the −1 *SD* cutoff and norms may not be broadly applicable (Johnson et al., 1999).

### NVIQ

Nonverbal general cognitive ability was assessed using the digital long form of the Raven’s Progressive Matrices-2^nd^ Ed. (Raven’s 2; *M* = 100, *SD* = 15) (Raven et al., 2018). As with language measures, administration followed test developer guidance for online administration (Pearson, 2023). The Raven’s 2 does not rely on language and is untimed, which enhances accessibility (Grondhuis et al., 2018). NVIQ was classified into bands for group comparisons: <75, 75 to 84, and ≥85. While prior work has used a cutoff as high as 80 on the Raven’s for low NVIQ (Silleresi, 2023), guidelines suggest 70 to 75 on cognitive tests is clinically significant (American Association on Intellectual and Developmental Disabilities, 2024). Note that the Raven’s alone does not convey intellectual disability, which requires comprehensive assessment (Raven et al., 2018).

### Sense of Community

Sense of community was measured using the Brief Sense of Community Scale (Peterson et al., 2008), which assesses psychological sense of community following an empirical model comprised of four constructs: needs fulfillment, group membership, influence, and emotional connection (McMillan & Chavis, 1986). This measure has been validated on racially and ethnically diverse youth and adults (ages 13-23+ years) from large, community-based samples that do not provide information on diagnoses (α = 0.92; Cardenas et al., 2021; Lardier Jr et al., 2018; Peterson et al., 2008). Respondents rate statements (1 = strongly disagree, 5 = strongly agree): (a) I can get what I need in this neighborhood, (b) This neighborhood helps me fulfill my needs, (c) I feel like a member of this neighborhood, (d) I belong in this neighborhood, (e) I have a say about what goes on in my neighborhood, (f) People in this neighborhood are good at influencing each another, (g) I feel connected to this neighborhood, and (h) I have a good bond with others in this neighborhood (Peterson et al., 2008). Item scores (a) and (b) are averaged to provide subscale scores for needs fulfillment (α = 0.80), (c) and (d) for group membership (α = 0.86), (e) and (f) for influence (α = 0.67), and (g) and (h) for emotional connection (α = 0.80), as well as an overall score. Higher scores indicate higher sense of community. Respondents completed the original measure for themselves or as a proxy. Caregivers-reported scores were significantly higher than self-reported scores, corresponding to “neutral to somewhat agree” and “somewhat disagree to agree”; see Supplementary Table 1.

### Unmet Service Needs and Barriers to Services

Unmet service needs and barriers to services were measured using survey items from the National Longitudinal Transition Study 2 (NLTS 2; Newman et al., 2011). The NLTS 2 followed a nationally representative sample of >11,000 adolescents (ages 13 to 18 years) receiving special education services from 2000 to 2010 and assessed services per federal special education legislation (“IDEIA,” 2004; Levine et al., 2007). Per prior work (Taylor & Henninger, 2015), this study adapted NLTS 2 items to assess unmet needs and asked if each service not received was needed. Respondents were self or caregiver and reported if: a) each of 16 services were received (psychological, speech-language, speech-language therapy or communication, career counseling or vocational/job skills training, personal assistant or in-home/in-classroom aide, medical services or diagnosis/evaluation related to special needs, occupation/life skills therapy or training, tutor, transportation, social work, assistive technology, respite care, reader or interpreter, physical therapy, orientation and mobility, audiology, other). All respondents reported if each of 12 items were barriers to having service needs met (cost, location, doctor or specialist does not accept insurance, not available, scheduling conflicts, ineligible, lack of information, transportation, quality, lack of time, language barrier, physical accessibility). Item scores were summed to provide totals of unmet service needs and of barriers. Caregiver-reported and self-reported unmet service needs and barriers did not significantly differ; see Supplementary Table 1.

### Communication Abilities

Social communication impairment was assessed using SRS-2 social communication impairment *t*-scores (Constantino, 2012). Group means of caregiver student report, caregiver adult report, and adult self-report forms did not significantly differ; see Supplementary Table 1. Functional communication was assessed using the Vineland Adaptive Behavior Scales-3^rd^ Ed. (VABS-3) domain-level form communication standard score (Sparrow et al., 2016). Caregivers indicate the frequency with which their child completes items. Item-level scores provide a standard score (*M* = 100, *SD* = 15), with higher scores indicating higher skills.

### Data Diagnostics and Analysis

Two trained research assistants independently scored and checked language and autism trait measures. The first author and research assistants discussed all disagreements until consensus was reached. NVIQ, functional communication, and social drivers of health measures were each auto-scored within their respective platforms. Next, data was examined for missingness. This study did not exclude any participant post-data collection. Missing data were minimal and came from different participants: (a) one missing language and NVIQ scores (did not complete assessment), (b) two missing SRS-2 scores (one for not completed, one form misplaced), and (c) two missing social drivers of health measures. Missing values (<5%) were imputed using predictive mean matching, a semi-parametric method appropriate for non-normal data, with one imputation (Little & Rubin, 2019). Data diagnostics and analyses took place in SPSS 29 (IBM Corp., 2023). Prior to analysis, data were checked for multicollinearity (VIF < 4, α = .05), linearity, and normality. Data were not transformed for normality except for the second research question, which *z*-scored and averaged PPVT-5, EVT-3, and SRT scores. To facilitate interpretation of model results, NVIQ was centered on 100 (Hoffman & Walters, 2022).

Forms for overall autism traits and social communication impairment *t*-scores were combined given similar effects across respondents. Despite small to medium effect sizes (Gaeta & Brydges, 2020), results were not statistically significant, and 95% confidence intervals were wide – and included zero – which indicates imprecision (Cohen, 1992; Correll et al., 2020; Lakens, 2013). Unmet service needs and barriers were also combined due to absence of significant results, wide 95% confidence intervals, small sample constraints, and unstable effect sizes (Kelley & Preacher, 2012). Though sense of community scores differed significantly by respondent, forms were combined to enable exploratory (versus inferential) interpretation and to address imbalance in group sizes (51 caregivers versus 20 self-report). This approach aligns with early-stage clinical research practices (De Los Reyes et al., 2013; Maxwell et al., 2018).

To assess whether LI status differentiated participants in communication scores and social drivers of health, Wald χ^2^ statistics tested for group differences, given NVIQ band, in social communication impairment scores, functional communication scores, overall sense of community score, number of unmet service needs, and number of barriers to services. Data were corrected for multiple comparisons using the Holm-Bonferroni method (Holm, 1979). Though not a primary outcome, a Fisher’s exact test assessed whether there was a significant association between LI status and NVIQ band, as well as descriptives (estimated marginal means, 95% confidence intervals, percentages). To address to what extent individual differences and social drivers of health contributed to concurrent communication scores, given limited sample size, Spearman (1904) correlations evaluated patterns between language, NVIQ, social drivers of health, and communication scores, with interpretation of was 0.25 as small, 0.40 as moderate, and 0.65 as large (Gaeta & Brydges, 2020). Correlations of ≥ 0.25 at an α < .05 were entered into regression models. As outcome variables were not normally distributed, separate generalized linear mixed effects models estimated the extent to which language *z*-scores, NVIQ centered on 100, and social drivers of health contributed to differences in functional communication scores and social communication impairment scores. Models were fit with GLMM procedures in SPSS 29 (IBM Corp., 2023), with fixed effects of predictors and by-participant random intercepts. Model fit was assessed using information criteria and likelihood ratio tests, with a α of .05.

## Results

### Participant Characteristics

Recruitment took place from 2022 to 2023, resulting in a sample of 73 participants (*M_age_* = 19.69, *SD* = 4.71, 13.27-30.47 years); see Tables 1 and 2. Over half the sample was in educational programming (*n* = 41, or 56.2%), with 21 (28.8%) in secondary education and 20 (27.4%) in post-secondary educational programs (e.g., day habilitation). Fisher’s exact test revealed an association between LI status and NVIQ band, *p* = .002, with six groups: 1) NVIQ < 75 without LI (*n* = 1); 2) NVIQ < 75 with LI (*n* = 10); 3) NVIQ of 75 to 84 without LI (*n* = 2); 4) NVIQ of 75 to 84 with LI (*n* = 8); 5) NVIQ ≥ 85 without LI (*n* = 29); and 6) NVIQ ≥ 85 with LI (*n* = 22). However, LI status and NVIQ band dissociated in over one-third of the sample. Three (4.11%) participants had NVIQ < 84 but not LI, and 22 (30.14%) had NVIQ ≥ 85 plus LI.

**Table 1.**
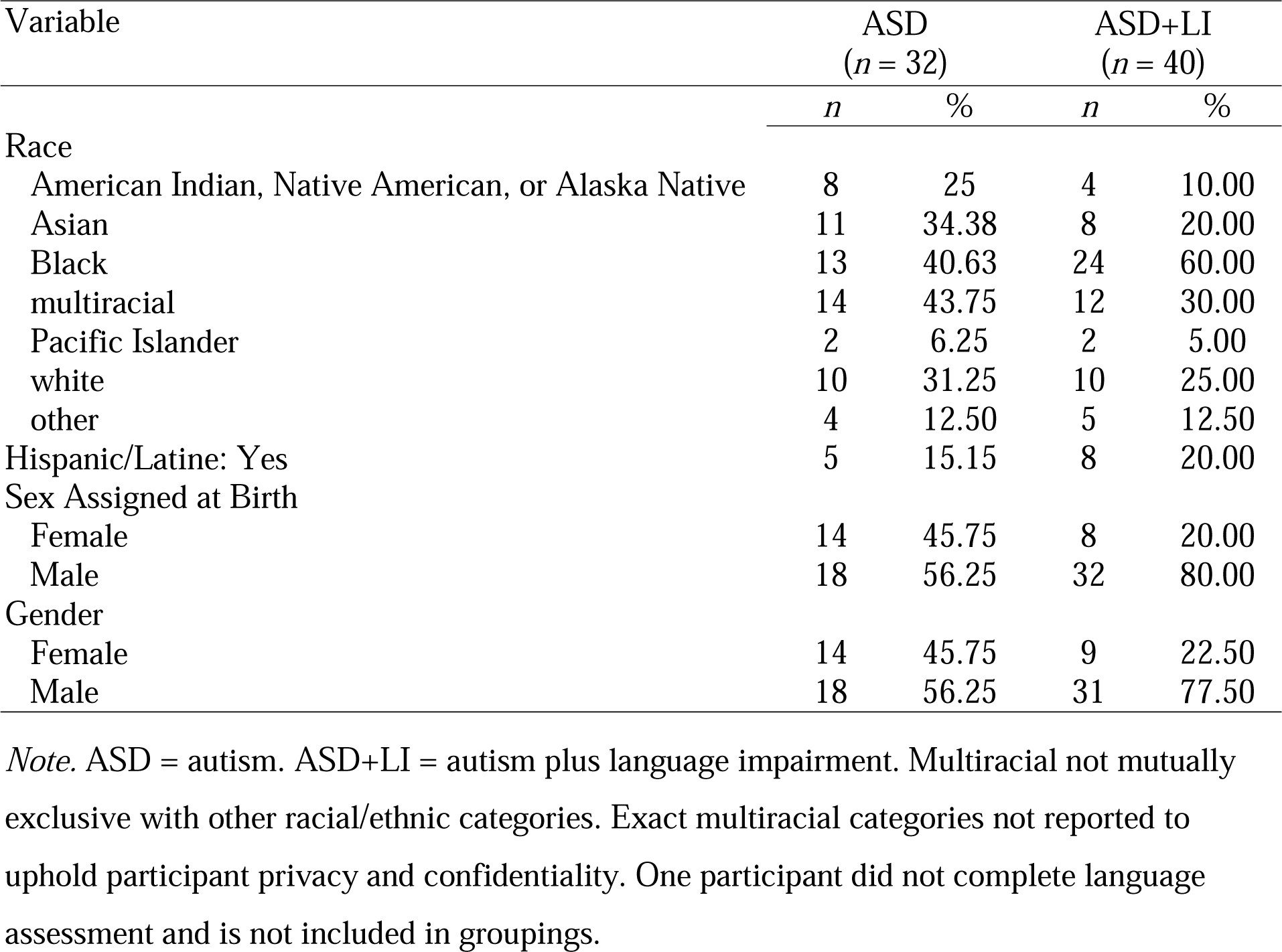
Participant Demo graphics.

| Variable | ASD<br>(*n* = 32) |  | ASD+LI<br>(*n* = 40) |  |
| --- | --- | --- | --- | --- |
|  | *n* | % | *n* | % |
| Race |  |  |  |  |
| American Indian, Native American, or Alaska Native | 8 | 25 | 4 | 10.00 |
| Asian | 11 | 34.38 | 8 | 20.00 |
| Black | 13 | 40.63 | 24 | 60.00 |
| multiracial | 14 | 43.75 | 12 | 30.00 |
| Pacific Islander | 2 | 6.25 | 2 | 5.00 |
| white | 10 | 31.25 | 10 | 25.00 |
| other | 4 | 12.50 | 5 | 12.50 |
| Hispanic/Latine: Yes | 5 | 15.15 | 8 | 20.00 |
| Sex Assigned at Birth |  |  |  |  |
| Female | 14 | 45.75 | 8 | 20.00 |
| Male | 18 | 56.25 | 32 | 80.00 |
| Gender |  |  |  |  |
| Female | 14 | 45.75 | 9 | 22.50 |
| Male | 18 | 56.25 | 31 | 77.50 |

*Note.* ASD = autism. ASD+LI = autism plus language impairment. Multiracial not mutually exclusive with other racial/ethnic categories. Exact multiracial categories not reported to uphold participant privacy and confidentiality. One participant did not complete language assessment and is not included in groupings.

**Table 2.** Participant Clinical Assessment Scores.

| Variable | ASD<br>(*n* = 32) |  |  | ASD+LI<br>(*n* = 40) |  |  |
| --- | --- | --- | --- | --- | --- | --- |
|  | *n* | *M* | *SD* | *n* | *M* | *SD* |
| NVIQ standard score | 32 | 99.00 | 13.90 | 40 | 84.03 | 15.00 |
| SRS-2 overall *t*-score | 32 | 73.94 | 11.81 | 38 | 71.11 | 11.10 |
| PPVT-5 receptive vocabulary standard score | 32 | 106.25 | 12.57 | 40 | 80.65 | 18.91 |
| EVT-3 expressive vocabulary standard score | 32 | 107.8 | 10.51 | 40 | 82.9 | 14.43 |
| SRT percent accuracy | 32 | 94.56 | 5.57 | 40 | 87.25 | 10.39 |
| CELF-5 Receptive Language Index | 32 | 99.41 | 10.54 | 40 | 68.16 | 15.31 |
| CELF-5 Expressive Language Index | 32 | 92.94 | 9.85 | 40 | 64.98 | 14.88 |

*Note.* ASD = autism. ASD+LI = autism plus language impairment. NVIQ assessed using Raven's 2 Progressive Matrices (Raven, 2018). SRS-2 = Social Responsiveness Scale-2nd Ed. (Constantino, 2012). PPVT-5 = Peabody Picture Vocabulary Test-5th Ed. (Dunn, 2019). EVT-3 = Expressive Vocabulary Test-3rd Ed. (Williams, 2019). SRT = Syllable Repetition Task (Shriberg et al., 2009). CELF-5 = Clinical Evaluation of Language Fundamentals-5th Ed. (Wiig et al., 2013). Scores replaced using single imputation of variable means for: CELF-5, PPVT-5, EVT-3, SRT, and NVIQ scores (*n* = 1), as well as SRS-2 total *t*-scores (*n* = 2). Twenty-five participants were >21 years.

### Communication Abilities and Social Drivers of Health by Language Impairment Status

Given NVIQ band, participants with and without LI did not significantly differ in communication scores, based on estimated marginal means (EMMs; see Table 3). However, clinical descriptions diverged. Functional communication scores were adequate for the autism group and moderately low for those with LI. Over three-fourths of participants with LI had low (29.4%) or moderately low (50%) functional communication scores compared to less than one-third of those without LI (29.4%; see Figure 1). Both groups showed moderate levels of social communication impairment, but distributions varied. Approximately two-thirds of participants with LI had moderate (32.5%) or severe levels of impairment (32.5%), whereas those without LI more often had severe impairment (48.5%) than moderate impairment (15.2%).

**Figure 1.**
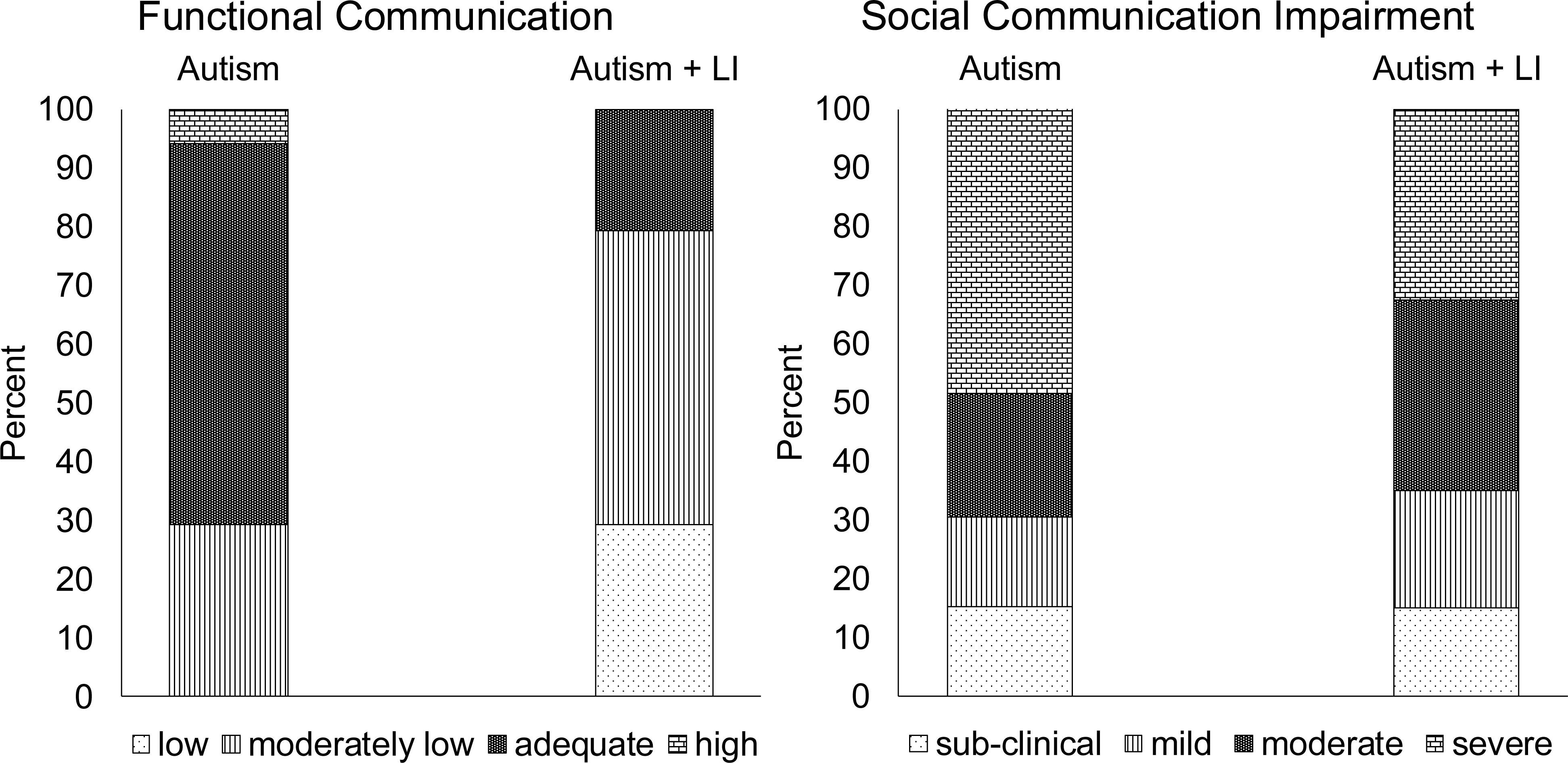
Communication abilities by group. LI = language impairment.

**Table 3.** Group Communication Scores and Social Drivers of Health (N = 73)

| Variable | Autism<br>(n = 33) | | Autism + LI<br>(n = 40) | | Wald $\chi^2$ df | | p |
| --- | --- | --- | --- | --- | --- | --- | --- |
|  | EMM | 95% CI | EMM | 95% CI |  |  |  |
| Unmet service needs | 3.31 | [2.28, 4.82] | 3.64 | [2.72, 4.87] | 0.16 | 1 | .693 |
| Barriers to services | 5.13 | [3.90, 6.74] | 6.01 | [5.01, 7.21] | 1.14 | 1 | .285 |
| Sense of community | 2.9 | [2.54, 3.26] | 3.04 | [2.72, 3.37] | 0.51 | 1 | .473 |
| SRS-2 SCI *t*-score | 74.1 | [69.34, 79.18] | 71.1 | [67.62, 74.63] | 1.26 | 1 | .261 |
| VABS-3 communication | 88.02 | [79.15, 97.89] | 76.3 | [71, 81.95] | 6.10 | 1 | .014 |

*Note.* LI = language impairment. EMM = estimated marginal means. Models fit with maximum likelihood estimation and robust covariance. Unmet service needs fit with negative binomial distribution and log link. Barriers to services fit with a Poisson distribution and log link. Sense of community and VABS-3 communication scores fit with Gaussian distribution and identity link. SRS-2 SCI *t*-scores fit with Tweedie distribution and identity link. Wald $\chi^2$ statistics and EMM control for NVIQ of < 75, 75 to 84, and ≥ 85. Data corrected for multiple comparisons using Holm-Bonferroni method, with an adjusted $\alpha$ of < .01 (Holm, 1979). Sense of community = Brief Sense of Community Scale overall score (Peterson et al., 2008). Unmet service needs and barriers to services = National Longitudinal Transition Survey-2 items (Newman et al., 2011), per Taylor & Henninger (2015). SRS-2 SCI = Social Responsiveness Scale-2nd Ed. social communication & interaction (Constantino, 2012).

LI status, given NVIQ, also did not significantly differentiate groups in social drivers of health (Table 3). However, clinical descriptions diverged. Both groups had a wide range of overall sense of community scores, with group EMMs corresponding approximately to “neutral.” Further, both groups reported varying unmet service needs (0-14) and barriers (0-11), with similar group EMMs of over three unmet service needs and five barriers. A majority of participants across groups reported receiving psychological or mental health services and medical services related to special needs; see Supplementary Table 2. Those with LI reported higher receipt of some services, such as personal assistant or in-class aide support (36.8% versus 6.8%) and speech-language therapy (47.4% versus 18.8%). Less common services received were orientation and mobility services, assistive technology, and respite care.

Among participants not receiving a given service, both groups most frequently reported unmet needs in career counseling or vocational/job skills training, occupation or life skills therapy or training, and speech-language therapy or communication services. Those with LI endorsed all services except for assistive technology as unmet needs at higher rates those without LI. Speech-language services were an unmet need for 50% of the LI group versus 38.5% of those without LI. For personal assistant or in-home/in-class aide support, the respective rates were 45.8% and 16.7%.

Endorsement of barriers to services was similarly high among both groups; see Supplementary Table 3. The most common barriers included the location of services, services not being available, providers not accepting insurance, ineligibility for services, cost, and scheduling conflicts; these were endorsed by a majority of participants with and without LI. Notably, those with LI endorsed poor service quality at over two times the rate of those without LI (57.9% versus 25%). Overall, LI status did not differentiate groups in communication scores or social drivers of health, but patterns suggest that those with LI may experience greater service gaps that are important for navigating daily life and communication.

### Model Results of Social Drivers of Health on Communication Scores

Next, analyses examined patterns between social drivers of health and communication scores; see Table 4. Spearman correlations showed functional communication was positively associated with language and NVIQ, and negatively associated with unmet service needs, social communication impairment, and service barriers. That is, higher functional communication scores corresponded with higher language and nonverbal cognitive ability, fewer service challenges, and lesser social communication impairment. Social communication impairment was negatively associated with both unmet service needs and sense of community, and positively associated with barriers to services. In other words, lower levels of impairment were linked to greater sense of community and fewer service-related challenges. Finally, higher language was associated with higher NVIQ and fewer unmet service needs. More unmet service needs and more barriers were associated with one another, as well as with lower sense of community. Overall, patterns suggest that language, NVIQ, and social drivers of health each relate to communication outcomes, but there was no one-to-one mapping.

**Table 4.**
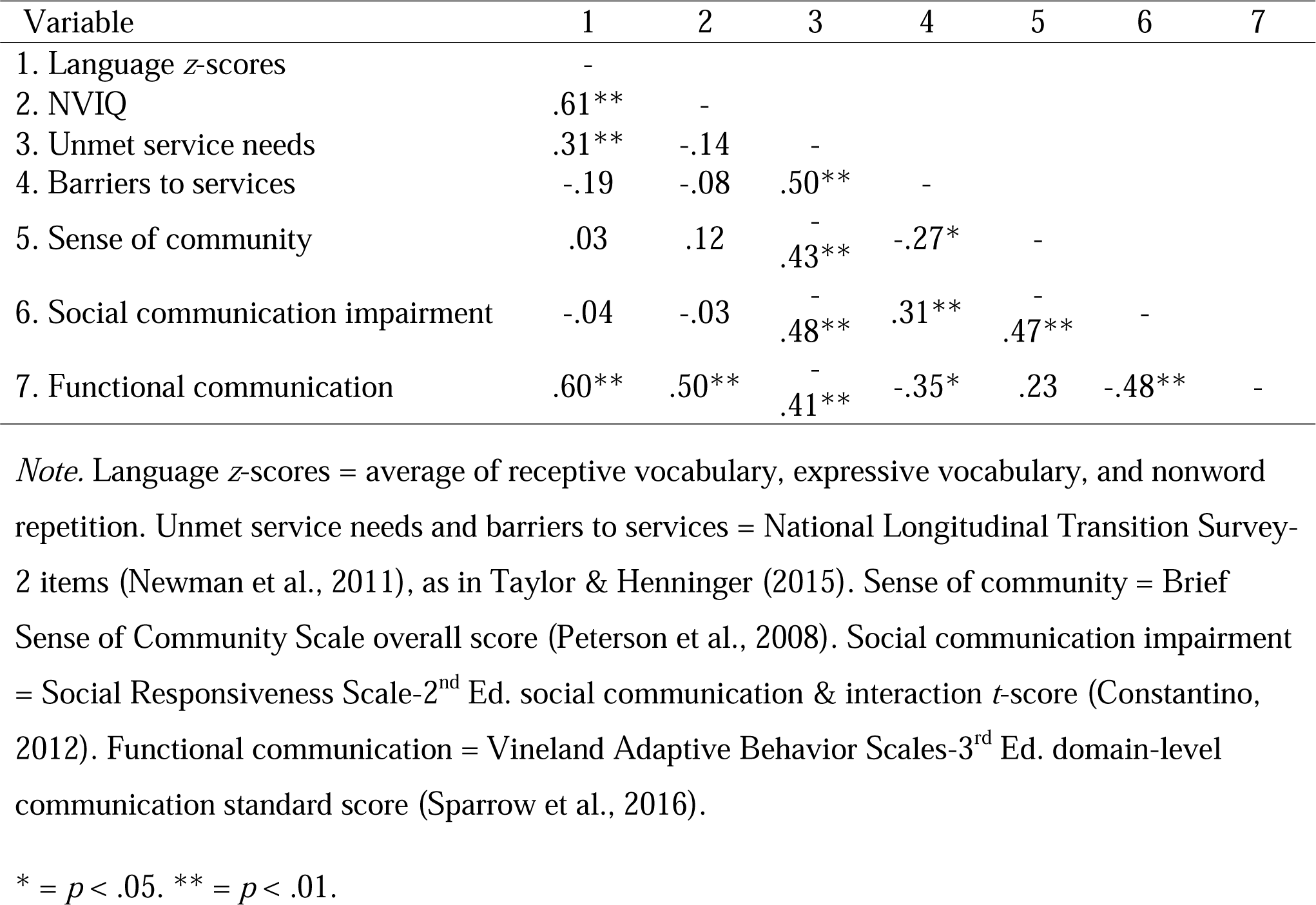
Spearman’s Correlations of Social Drivers of Health, Language, NVIQ, and Communication Scores.

Generalized linear mixed model results showed that language scores, but not NVIQ, unmet service needs, or barriers, significantly contributed to differences in functional communication scores; see Table 5 and Figure 2. Baseline variability was significant, τ^2^ = 151.06, *z* = 2.55, *p* = .012. Including language, NVIQ (centered on 100), unmet service needs, and barriers to services improved model fit and explained 40.5% of the variance. However, interindividual variability remained significant, τ^2^ = 94.38, *z* = 2.40, *p* = .016. The expected score at baseline (language *z*-score = zero, NVIQ = 100, no unmet service needs or barriers) was 95.08, or an “adequate” functional communication level. Each one-unit increase in language *z*-scores was associated with a 5.54-point increase in functional communication scores.

**Figure 2.**
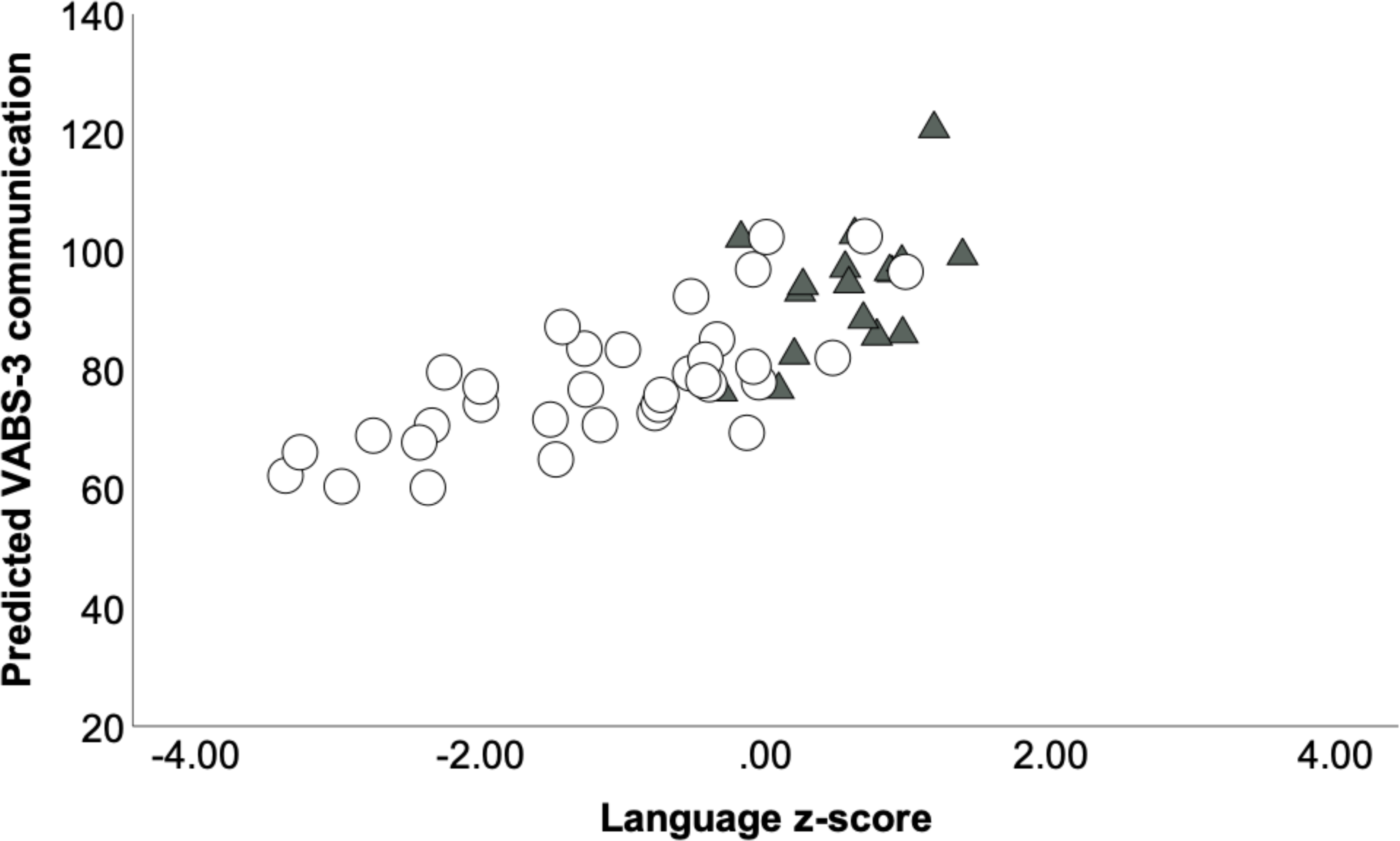
Observed language z-scores by Vineland Adaptive Behavior Scales-3^rd^ Ed. communication standard scores, based on model results including language z-scores, NVIQ, unmet service needs, and barriers to services. Circles = autism plus language impairment. Triangles = autism without language impairment. Language z-scores = average of receptive vocabulary, expressive vocabulary, and nonword repetition accuracy.

**Table 5.** Model Results of Functional Communication Scores.

| Parameter | Model 1 |  |  | Model 2 |  |  |
| --- | --- | --- | --- | --- | --- | --- |
| | $\beta$ | *SE* | 95% CI | $\beta$ | *SE* | 95% CI |
| Intercept | 83.20** | 2.41 | [78.36, 88.04] | 95.08** | 4.12 | [86.79, 103.37] |
| Language *z*-scores |  |  |  | 5.44** | 1.67 | [1.94, 8.93] |
| NVIQ centered on 100 |  |  |  | .16 | .11 | [-.07, .40] |
| Unmet service needs |  |  |  | -.65 | .66 | [-2.00, .71] |
| Barriers to services |  |  |  | -.96 | .69 | [-2.34, .43] |
| AIC | 435.62 |  |  | 405.44 |  |  |
| BIC | 439.19 |  |  | 408.82 |  |  |
| Marginal pseudo-$R^2$ | .000 | | | .405 | | |
| Conditional pseudo-$R^2$ | .500 | | | .702 | | |
| -2 LL | 431.37 |  |  | 401.16 |  |  |
| *df* | 2 |  |  | 6 |  |  |
| LRT |  |  |  | 30.21*** |  |  |

*Note.* Functional communication scores = Vineland Adaptive Behavior Scales-3 communication domain standard scores (Sparrow et al., 2016). Language *z*-scores = average of receptive vocabulary, expressive vocabulary, and nonword repetition. Unmet service needs and barriers to services = National Longitudinal Transition Survey-2 items (Newman et al., 2011), as in Taylor & Henninger (2015). Models fit with a Gaussian distribution and identity link function, with a random intercept, variance components structure, Satterthwaite approximation, and robust covariance estimation. LRT = likelihood ratio test, using a chi-square statistic with 4 degrees of freedom.
\* = $p < .05$ . \*\* $p < .01$ . \*\*\* $p < .0001$ .

In contrast, in the regression model, sense of community was significantly associated with social communication impairment, whereas unmet service needs and barriers to services were not; see Table 6 and Figure 3. There was significant baseline variability, τ^2^ = 61.65, *z* = 3.04, *p* = .002. Adding sense of community, unmet service needs, and barriers to services improved model fit and accounted for 27.3% of the variance in impairment scores. At baseline (no unmet service needs of barriers), the expected *t*-score was 79.50, indicating a “high” level of social communication impairment. Each one-unit increase in sense of community was associated with a 4.18-point decrease in impairment scores. Individual variability remained significant in the final model, τ^2^ = 46.21, *z* = 2.94, *p* = .003.

**Figure 3.**
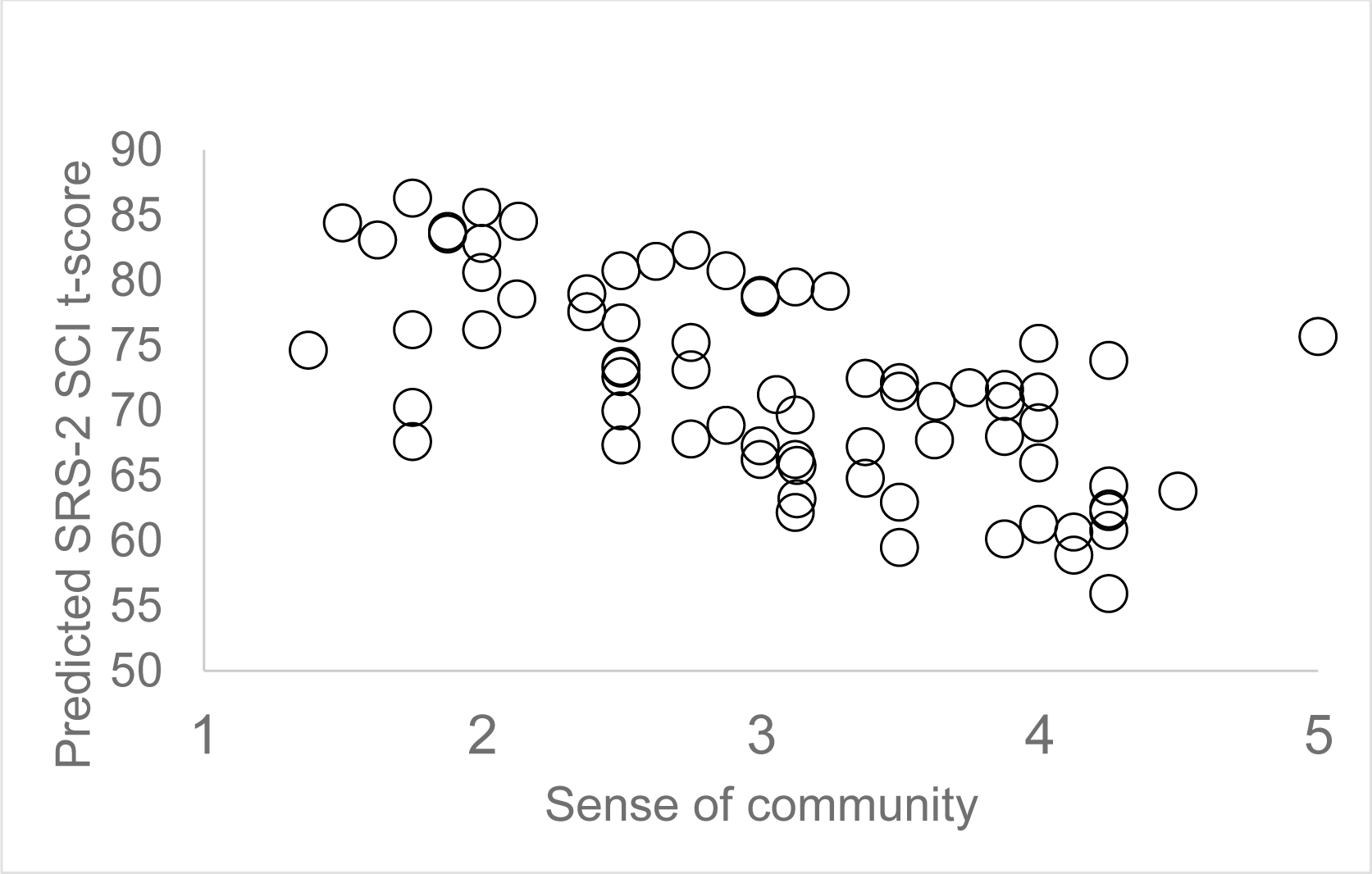
Observed sense of community by Social Responsiveness Scale-2^nd^ Ed. Social communication impairment t-score, based on model results including sense of community, unmet service needs, and barriers to services.

**Table 6.**
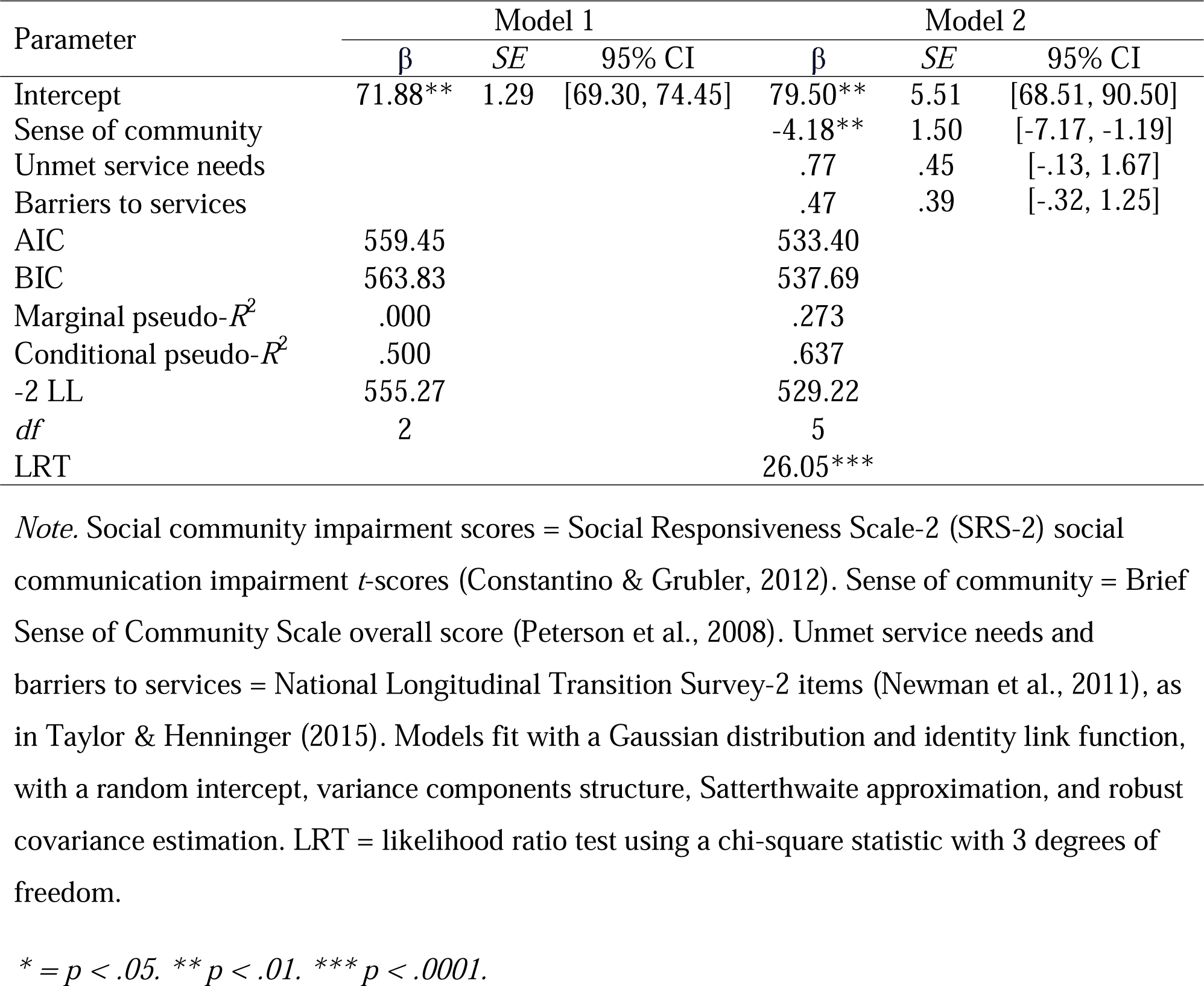
Model Results of Social Communication Impairment Scores.

## Discussion

This report is a first step in understanding social drivers of health in minoritized autistic adolescents and young adults ranging in language skills. Findings are consistent with prior reports on functional communication in adults with LI (Johnson et al., 1999), as well as on the relevance of community supportiveness (Devenish et al., 2020; Talò et al., 2014). In this study, communication and social drivers of health were variable. These results are important for understanding linguistic heterogeneity in this population. Findings lay groundwork for better understanding the transition to adulthood (Magiati et al., 2014; Schott et al., 2022).

### Measurement Issues: Language Impairment and Communication

Calls for multi-domain assessment of language in autism have grown, as over-relying on individual measures leads to low clinical validity (Schaeffer et al., 2023). Criteria for LI in this study spanned semantics, morphology, syntax, and phonology. This approach, consistent with prior work (Johnson et al., 1999), avoids reliance on cognitive referencing and reflects the fact that language and NVIQ can dissociate in autism (Manenti et al., 2024; Silleresi et al., 2020; WHO, 2022).

Still, limitations remain. No comprehensive normed expressive-receptive language tools exist for adults over age 21 years, which affects classification reliability (e.g., Fidler et al., 2011; Johnson et al., 1999). Observational tools, such as natural language sampling, may offer additional insight (Butler et al., 2023), but were not feasible in this study. Especially with a sample that is historically marginalized in research, time is a constraint. Thus, this study balanced duration of activities with administration of behavioral assessment and questionnaires.

Findings also have implications for conceptualizing communication. Communication is variably defined and may refer to verbally expressing needs, managing social situations, or self-perceived abilities (Cummins et al., 2020; Riglin et al., 2021; Sterrett et al., 2024)—and does not necessarily consider structural language skills. Here, more participants without LI had adequate functional communication, which is consistent with prior work on adults with LI (Johnson et al., 1999), but groups did not differ by LI status in communication scores. These mixed findings demonstrate how over-reliance on measures may mask variation.

Overall, findings emphasize the trade-offs in approaches to measurement. Broad classification (e.g., LI) can obscure nuance, while narrowly focused tools may miss important variability. While there is no consensus on LI or communication, measurement should be guided by clear goals and an understanding of how data will be used (Messick, 1990).

### Patterns Between Social Drivers of Health and Communication Abilities

Communication scores were associated with individual differences in language and sense of community, but not services or NVIQ. As this was a cross-sectional analysis, the observed associations do not imply causality or directionality. While higher sense of community corresponded with lower social communication impairment, it is also plausible that greater impairment limits access or participation in community settings, reducing their sense of community. Sense of community was lower in this sample than in primarily white adults from the general population (2.9-3.04 versus 3.81; Peterson et al., 2008). Though beyond the scope of this study, research on Black and Hispanic youth and adults suggests these lower scores may reflect social marginalization (Cardenas et al., 2021; Davis et al., 2024; Scott et al., 2021).

This study included social drivers of health, language, and NVIQ to avoid collapsing heterogeneity among minoritized autistic adolescents and young adults (Bronfenbrenner, 1977; Plaut, 2010). However, interpreting heterogeneity is complex. Language scores corresponded with, but did not contribute to, functional communication scores when accounting for NVIQ< unset service needs, and barriers. Prior work found 12.8% of autistic and nonautistic adults with a history of LI reported “normal speech and language functioning” (Johnson et al., 1999), and links between cognitive abilities and functional communication in autism remain confounded by use of full-scale IQ (Gronshuis et al., 2018; Matthews et al., 2015). Additionally, counts of unmet service needs and barriers may miss nuanced differences within and between individuals (Burke et al., 2024).

Community support may play a role in supporting self-expression and communication during the transition to adulthood (Farley et al., 2009). Social communication impairment more directly aligned with expected results, according to theory (Bronfenbrenner, 1977) and prior work on adults from the general population (Talò et al., 2014). For autistic adults, peer understanding may affect social interaction more than individual traits, such as social motivation or cognition (Crompton et al., 2020a; Morrison et al., 2020a). Hence, higher impairment could arise from external factors that reduce community access and lower perceived sense of belonging (Cameron et al., 2022). Finally, lower sense of community in this sample may reflect intersecting experiences of race and disability (Bolick, 2008; Littman, 2022). Individuals may opt out of communication experiences when environments do not offer supports and feel exclusionary. However, the dynamics between agency and structural constraints are unknown.

### Implications for Autistic Individuals and Community Members

Though there is growing awareness of heterogeneity in language in autism (Schaeffer et al., 2023) and of the transition to adulthood (Roux et al., 2015), documenting this variability is critical for advocacy. Rather than rely on limited information–which risks reinforcing stereotypes–developing a nuanced understanding of autistic adolescents’ and young adults’ profiles is essential to recognizing them as whole individuals (Kover & Abbeduto, 2023). Amid ongoing emphasis on individual traits over social drivers of health in research on transition-aged autistic individuals (Anderson et al., 2018a; Magiati et al., 2014), attending to environmental factors is key to understanding autism across the lifespan (Schendel et al., 2022). Doing so is prerequisite to enhancing person-environment fit (Lai et al., 2020), align supports with individual goals, and to shift intervention toward community-level change rather than placing the onus solely on autistic individuals. Findings from this study also underscore the value of using appropriate conceptual approaches for diversity (Plaut, 2010). While identifying disparities is relevant for developing targeted interventions, white-minoritized comparisons require clear justification.

## Limitations

This study had several limitations. The sample was small and included transition-aged individuals who used primarily language to communicate (Hughes et al., 2023). This study was cross-sectional, limiting the ability to examine directionality or causality among observed relationships, which is relevant as service needs may evolve across early to later adulthood (Schendel et al., 2022). Also, although this study was motivated by a social-ecological model (Bronfenbrenner, 1977), analysis did not examine bidirectional patterns between social drivers of health and communication abilities. These limit the generalizability of findings. Different respondents also completed questionnaires, which may have contributed to the observed differences in sense of community; without robust sampling and replication, determining whether effects are “true” is futile (De Los Reyes et al., 2013). Moreover, including qualitative measures to elucidate how the intersection of race with disability shapes experiences (Annamma et al., 2013), such as sense of community and services access would strengthen interpretation of the data. Given concerns of feasibility, this study did not comprehensively assess multi-level, multi-domain social drivers of health that shape the communication environment (National Institute on Minority Health and Health Disparities, 2017), or clinical traits, such as mental health conditions (Kraper et al., 2017). Finally, the clinical and real-world utility of communication abilities, as assessed in this study, are unknown.

## Future Directions

Findings and limitations highlight priorities for future research. Social drivers of health measures used in this study may differ from those used in clinical settings (e.g., economic stability; Sokol et al., 2019), and understanding how these intersect is essential for improving outcomes in autism (Schendel et al., 2022). Although prior work documents healthcare barriers for autistic adults, such as patient-provider communication challenges, sensory sensitivities, and mismatches in care delivery, there is little attention to the roles of racial or ethnic minoritization or language (Mason et al., 2019). Similar gaps exist in tailoring mental and physical healthcare delivery for autistic adults (ages 18-77 years), including clinician knowledge and environmental factors (Brice et al., 2021). Addressing these challenges requires robust sampling and longitudinal data to examine how language, communication, and social drivers of health interact over time. Future work should integrate clinical communication assessments with person-centered measures, as both internal and contextual factors shape communication in autistic adults (Cummins et al., 2020). Enhancing the ecological validity of measuring social drivers of health – by focusing on meaningful benefits and outcomes of assessment – is key to reducing support gaps in the transition to adulthood. Given environmental barriers and limited provider preparation to interact with autistic adults (Doherty et al., 2022), it is critical to incorporate perspectives from autistic adolescents and young adults, caregivers, providers, and organizations to generate research for change (Field et al., 2014).

## Conclusions

In examining a minoritized sample of autistic adolescents and young adults ranging in language skills and NVIQ, this cross-sectional study documented relationships between social drivers of health and communication abilities. Patterns between language and functional communication, as well as sense of community and social communication impairment, support a model of communication outcomes as arising from individual-environment interactions. Findings emphasize the importance of the environment in shaping communication for autistic individuals during the transition to adulthood, as well as of a holistic approach to assessment and services.

## Data Availability Statement

Data for this manuscript are unavailable due to ethical reasons. Participants or caregivers (whoever provided informed consent) completed a graded consent form modeled after FluencyBank, which allows them choice in what data to share with the research community: none, de-identified data, transcripts, and/or audio-recordings. They opted to not share their data.

## Contributor Roles Taxonomy Statement

**TG** (conceptualization, methodology, formal analysis, data curation, writing – original draft, visualization, project administration, supervision), **AE** (methodology, formal analysis, data curation, writing – review & editing), **LB** (methodology, formal analysis, writing – review & editing), **CL** (conceptualization, methodology, formal analysis, writing – review & editing), **IC** (conceptualization, methodology, writing – review & editing), **KGP** (conceptualization, methodology, writing – review & editing).

## Acknowledgements

TG was supported by an American Speech-Language-Hearing Foundation New Investigators Research Grant (PI: Girolamo), NIDCD L70 DC021323 (PI: Girolamo), NIDCD T32 DC001703 (PI: Eigsti), and NIDCD R21 DC021769-01A1 (PI: Girolamo).

