## Supplemental Tables for "The role of social drivers of health in communication abilities of autistic adolescents and young adults"

| **Supplementary Table 1**  *Independent Sample T-Test and One-Way Analysis of Variance Results of Differences in Scores by Respondent* | | | | | | | | | | | | | | | | | | | | | |
| --- | --- | --- | --- | --- | --- | --- | --- | --- | --- | --- | --- | --- | --- | --- | --- | --- | --- | --- | --- | --- | --- |
| Measure | Self | | | Caregiver | | | |  | |  | |  | | *t(69*) | | *p* | | Cohen's *d* | | 95% CI | |
|  | *n* | *M* | *SD* | | *n* | *M* | *SD* | |  | |  | |  | |  | |  | |  | |  |
| BSCS overall score | 20 | 2.51 | 0.63 | | 51 | 3.27 | 0.86 | |  | |  | |  | | -3.66 | | < .001 | | 0.81 | | -1.50, -0.42 |
| Total unmet service needs | 20 | 3.12 | 3.15 | | 51 | 3.9 | 3.51 | |  | |  | |  | | 0.91 | | .365 | | 3.25 | | -2.79, .758 |
| Total barriers to services | 20 | 5.65 | 3.57 | | 51 | 5.47 | 3.31 | |  | |  | |  | | 0.2 | | .841 | | 3.38 | | -.464, .570 |
|  | Self - Adult | | | Caregiver - Student | | | | Caregiver - Adult | | | | | | *F*(2,68) | | *p* | | partial ε² | | 95% CI | |
|  | *n* | *M* | *SD* | | *n* | *M* | *SD* | | *n* | | *M* | | *SD* | |  | |  | |  | |  |
| SRS-2 overall *t*-score | 20 | 76.15 | 8.7 | | 37 | 70.68 | 11.86 | | 14 | | 69.07 | | 11.93 | | 2.98 | | .058 | | .05 | | -.029, .182 |
| SRS-2 SCI *t*-score | 20 | 77.8 | 8.45 | | 37 | 70.92 | 12.12 | | 14 | | 69.86 | | 12.18 | | 2.15 | | .124 | | .03 | | -.029, .151 |
| *Note.* BSCS = Brief Sense of Community Scale (Peterson et al., 2008). SRS-2 = Social Responsiveness Scale-2 (Constantino, 2012). SCI = social communication impairment. | | | | | | | | | | | | | | | | | | | | | |

| **Supplementary Table 2**  *Descriptive Frequencies of Services Received and Unmet Service Needs* | | | | | | | | | | | | |
| --- | --- | --- | --- | --- | --- | --- | --- | --- | --- | --- | --- | --- |
| Service | Autism (*n* = 32) | | | | | | Autism+LI (*n* = 38) | | | | | |
|  | Received | | Unmet | | Not Needed | | Received | | Unmet | | Not Needed | |
|  | *n* | % | *n* | % | *n* | % | *n* | % | *n* | % | *n* | % |
| Audiology services | 2 | 6.25 | 1 | 3.33 | 29 | 96.67 | 3 | 7.89 | 5 | 14.29 | 30 | 85.71 |
| Assistive technology services/devices | 2 | 6.25 | 7 | 23.33 | 23 | 76.67 | 8 | 21.05 | 4 | 13.33 | 26 | 86.67 |
| Career counseling or vocational/job skills training | 5 | 15.63 | 15 | 55.56 | 12 | 44.44 | 16 | 42.11 | 16 | 72.73 | 6 | 27.27 |
| Medical services or diagnosis/evaluation related to special needs | 16 | 50.00 | 2 | 12.50 | 14 | 87.50 | 22 | 57.89 | 6 | 37.50 | 10 | 62.50 |
| Occupation/life skills therapy or training | 6 | 18.75 | 10 | 38.46 | 16 | 61.54 | 15 | 39.47 | 12 | 52.17 | 11 | 47.83 |
| Orientation and mobility services | 0 | 0.00 | 2 | 6.25 | 30 | 93.75 | 2 | 5.26 | 4 | 11.11 | 32 | 88.89 |
| Other services | 4 | 12.50 | 6 | 21.43 | 22 | 78.57 | 6 | 15.79 | 11 | 34.38 | 21 | 65.63 |
| Personal assistant or in-home/in-classroom aide | 2 | 6.25 | 5 | 16.67 | 25 | 83.33 | 14 | 36.84 | 11 | 45.83 | 13 | 54.17 |
| Physical therapy | 5 | 15.63 | 3 | 11.11 | 24 | 88.89 | 4 | 10.53 | 5 | 14.71 | 29 | 85.29 |
| Psychological/mental health services or counseling | 25 | 78.13 | 2 | 28.57 | 5 | 71.43 | 21 | 55.26 | 7 | 41.18 | 10 | 58.82 |
| Reader or interpreter | 0 | 0.00 | 4 | 12.50 | 28 | 87.50 | 2 | 5.26 | 7 | 19.44 | 29 | 80.56 |
| Respite care | 1 | 3.13 | 4 | 12.90 | 27 | 87.10 | 10 | 26.32 | 12 | 42.86 | 16 | 57.14 |
| Social work service | 3 | 9.37 | 9 | 31.03 | 20 | 68.97 | 13 | 34.21 | 10 | 40.00 | 15 | 60.00 |
| Speech-language therapy or communication services | 6 | 18.75 | 10 | 38.46 | 16 | 61.54 | 18 | 47.37 | 10 | 50.00 | 10 | 50.00 |
| Transportation services | 3 | 9.37 | 11 | 37.93 | 18 | 62.07 | 17 | 44.74 | 9 | 42.86 | 12 | 57.14 |
| Tutor | 4 | 12.50 | 6 | 21.43 | 22 | 78.57 | 10 | 26.32 | 12 | 42.86 | 16 | 57.14 |
| *Note.* Services organized alphabetically. Percentages for unmet service needs are calculated based on respondents who saw each item (i.e., did not receive a given service). Not needed = service not received and not needed. | | | | | | | | | | | | |

| **Supplementary Table 3**  *Descriptive Frequencies of Endorsement of Individual Barriers to Services* | | | | |
| --- | --- | --- | --- | --- |
|  | Autism  (*n* = 32) | | Autism+LI  (*n* = 38) | |
|  | *n* | % | *n* | % |
| Cost of services | 19 | 59.38 | 21 | 55.26 |
| Doctor or specialist does not accept insurance | 19 | 59.38 | 22 | 57.89 |
| Getting information about services | 15 | 46.88 | 24 | 63.16 |
| Lack of time for services | 14 | 43.75 | 16 | 42.11 |
| Language barrier | 0 | 0.00 | 1 | 2.63 |
| Not eligible for services | 18 | 56.25 | 22 | 57.89 |
| Physical accessibility | 2 | 6.25 | 3 | 7.89 |
| Poor service quality | 8 | 25.00 | 22 | 57.89 |
| Scheduling conflict | 15 | 46.88 | 20 | 52.63 |
| Services not available | 19 | 59.38 | 28 | 73.68 |
| Transportation | 11 | 34.38 | 19 | 50.00 |
| Where services are provided (e.g., geographic distance) | 20 | 62.50 | 29 | 76.32 |
| *Note.* Barriers organized alphabetically. One participant did not complete language measures and is not grouped. Two participants did not complete services questionnaires. | | | | |
